# The impact of early scientific literature in response to COVID-19: a scientometric perspective

**DOI:** 10.1101/2020.04.15.20066183

**Authors:** Davide Golinelli, Andrea Giovanni Nuzzolese, Erik Boetto, Flavia Rallo, Manfredi Greco, Fabrizio Toscano, Maria Pia Fantini

## Abstract

**Background:** In the early phases of a new pandemic, identifying the most relevant evidence and quantifying which studies are shared the most can help researchers and policy makers. The aim of this study is to describe and quantify the impact of early scientific production in response to COVID-19 pandemic.

**Methods:** The study consisted of: 1) review of the scientific literature produced in the first 30 days since the first COVID-19 paper was published; 2) analysis of papers’ metrics with the construction of a “Computed-Impact-Score” (CIS) that represents a unifying score over heterogeneous bibliometric indicators. In this study we use metrics and alternative metrics collected into five separate categories. On top of those categories we compute the CIS. Highest CIS papers are further analyzed.

**Results:** 239 papers have been included in the study. The mean of citations, mentions and social media interactions resulted in 1.63, 10 and 1250, respectively. The paper with highest CIS resulted “*Clinical features of patients*[…]” by Chaolin Huang et al., which rated first also in citations and mentions. This is the first paper describing patients affected by the new disease and reporting data that are clearly of great interest to both the scientific community and the general population.

**Conclusions:** The early response of scientific literature during an epidemic does not follow a pre-established pattern. Being able to monitor how communications spread from the scientific world toward the general population using both traditional and alternative metric measures is essential, especially in the early stages of a pandemic.

## 1. Background

Demographic evolution, removal of geographical barriers and climate change are causing both a shift in the endemic areas of many infectious diseases and the onset of new pathogens, with a progressive and parallel increase in the risk of epidemics and pandemics (van Doorn 2014; Baylis 2017).

Coronaviruses are highly prevalent in many species and are susceptible to frequent recombination and mutations of their genomes. On 31 December 2019, the World Health Organization (WHO) China Country Office was informed of cases of pneumonia of unknown etiology detected in Wuhan City (Hubei Province, China), possibly associated with exposures in a seafood wholesale market in the same city (WHO 2020). The cause of the syndrome was a new type of coronavirus isolated on 7th January 2020 and named Severe Acute Respiratory Syndrome CoronaVirus 2 (SARS-CoV-2). Formerly known as the 2019 novel coronavirus (2019-nCoV), SARS-CoV-2 is a positive-sense single-stranded RNA virus that is contagious among humans and is the cause of the coronavirus disease 2019, hereinafter referred to as COVID-19 (CDC 2020; Gorbalenya 2020; WHO 2020). The symptoms of COVID-19 infection appear after an incubation period of approximately five days and include fever, cough, and fatigue, sputum production, headache, haemoptysis, diarrhoea, dyspnea, and lymphopenia (Rothana 2020). While most people with COVID-19 develop only mild or uncomplicated illness, approximately 14% develop severe disease that requires hospitalization and oxygen support, and 5% require admission to an intensive care unit. In severe cases, COVID-19 can be complicated by the acute respiratory distress syndrome (ARDS), sepsis and septic shock, multiorgan failure, including acute kidney injury and cardiac injury (Yang 2020).

On January 30, 2020, the International Health Regulations Emergency Committee of the World Health Organization (WHO) appointed the COVID-19 outbreak a “public health emergency of international concern” (PHEIC). On March 11th 2020 WHO declared COVID-19 is pandemic (MacLaren 2020; Wang 2020; WHO 2020).

Worldwide, COVID-19 is showing critical issues in the response of health systems, which are put to the test by this emergency. Together with the fast spread of a new pathogen, we are witnessing a relatively new phenomenon, defined as “infodemic” by the WHO (WHO, 2020).

An “infodemic” represents the uncontrolled spread of false or “exaggerated” information (i.e. misinformation) relating to the pathogen or epidemic, which might determine an unpredictable response by the population. This response can translate into an increase in the concern of the population towards the epidemics, and even outright panic. This can also lead to public unrest or other consequences that can be difficult to control. Misinformation, like a virus itself, can be easily transmitted from person to person. The WHO has therefore highlighted that the spread of unconfirmed or incorrect information can be very dangerous for public health^1^.

The information relating to infectious outbreaks have a complex dynamic and occur at various levels. On the one hand, the scientific community is quickly activated on producing evidence, studies and articles that describe the new pathogen, the first cases, the methods of transmission, etc. For example, the first scientific article related to COVID-19 was indexed on Pubmed on January 14th 2020 (Bagoch 2020). In it, authors reported that a cluster of pneumonia of unknown aetiology was published on ProMED-mail, possibly related to contact with the Huanan seafood wholesale market in Wuhan, China, and warned for the potential international spread via commercial air travel.

On the other hand, the communication channels of the official sources (WHO, Ministries of Health, etc.) are committed to collecting, filtering and transmitting true and confirmed information, in order to provide a public service and to contain the population response.

In turn, traditional media (newspapers, periodicals, etc.), both digital and printed, resume the news and disseminate it. At this level, misinformation can occur. That is, the creation - more or less fraudulent - of false news that can determine an emotional response in the population, creating false beliefs or panic.

The last level of communication is on social media (e.g. Twitter, Facebook, Instagram, etc.), where individual citizens can share news and messages, communicating their feelings and their point of view on the subject.

These levels of communication are interconnected. For example, the sudden onset of a new virus forces the scientific community to describe the index-case, by publishing a paper in a scientific journal. Subsequently, if it deems it necessary, the Ministry of Foreign Affairs of one Country can indicate the risk for people travelling to the area where the case occurred. A newspaper can later resume the news, which can be shared and commented by individual users on social media. In this framework, misinformation might be the most dangerous and contagious aspect, as underlined by the WHO. Similarly to disease outbreak analysis, a viral content on the internet can also be seen as a chain reaction. Therefore, as misinformation can be considered as a public health threat, in the early stages of a pandemic it is important to contain false information and to disseminate correct data that may come primarily from scientific studies. At the present, it is essential that methodologically solid information is disseminated, both to avoid misinformation and to spread only real world evidence, possibly through peer-reviewed articles. This despite the fact that the peer review at this stage is done less rigorously because of the emergency. It is important that the information gaps are filled, but it is also important to contrast the infodemic with solid information, news and data.

Summing up, in the early phases of a new pandemic, the scientific and academic community is quickly activated on producing evidence and scientific articles. However, traditional media and social networks resume and disseminate information in a proper or inappropriate way. Identifying the most relevant evidence produced by the scientific community and quantifying which studies and which data are shared the most in the world can help researchers and policymakers in focusing on the most relevant ones and controlling the epidemic. This can be done by capturing and measuring traditional citations of scientific papers but also through the use of innovative and alternative scientometrics tools. Scientometrics is the field of study which measures and analyses scientific literature, including the measurement of the impact of research papers and the use of such measurements in policy and management contexts. Alternative metrics (a.k.a. “altmetrics”) are gaining increasing interest in the scientometrics community as they can capture both the volume and quality of attention that a research work receives online. Altmetrics are non-traditional research impact measures that are based on web-based environments. Altmetrics measurement derives from the social web and is increasingly used as an early indicator of research impact (P. Wouters et al. 2015; J. Ravenscroft et al. 2017; L. Bornmann, R. Haunschild 2018; Nuzzolese et al. 2019).

The aim of this study is to describe and quantify the impact - in terms of dissemination of knowledge - of early scientific production in response to the COVID-19 pandemic.

## 2. Methods

We conducted a twofold study which includes a review of the early scientific literature and a scientometric analysis. Specifically, the study consisted of the two following phases: 1) review of the scientific literature produced in the first 30 days since the first COVID-19 paper was published on MEDLINE/Pubmed; 2) identification of the Digital Object Identifiers (DOI) for each paper and analysis of citations and metrics measures to quantify their communicative impact (i.e. scientometric analysis).

### 2.1 Scientific literature review

The initial search was implemented on February 20, 2020 in MEDLINE/Pubmed. The search query consisted of terms considered by the authors to describe the new epidemic: [coronavirus* OR Pneumonia of Unknown Etiology OR Covid-19 OR nCoV]. Although the virus name was updated to SARS-CoV-2 by the International Committee on Taxonomy of Viruses on February 11th 2020 (Gorbalenya 2020), we performed the search using the term “nCoV” because it was presumed that no one, between February 11 and 13, would have used the term “SARS-COV-2”. To achieve the highest sensitivity, we decided to use only a combination of keywords avoiding Mesh terms. Asterisks are used to truncate words, so that every ending after the asterisks was searched. We placed a language restriction for English, without other limits.

Furthermore, we limited the search to the following time-span: from December 1, 2019 to February 13, 2020. Due to the extraordinary rapidity with which scientific papers have been electronically published online (i.e. ePub), it may happen that some of these have indicated a date later than February 13 2020 as publication date.

A two-stage screening process was used to assess the relevance of identified studies. For the first level of screening, only the title and abstract were reviewed to preclude waste of resources in procuring articles that did not meet the minimum inclusion criteria. Titles and abstracts of studies initially identified were checked by two independent investigators (E.B. and F.R.).

All citations deemed relevant after title and abstract screening were procured for subsequent review of the full-text article. A form was developed to extract study characteristics such as publication date, publication type, aim of the study, and authors’ nationality.

### 2.2 Scientometric analysis

In order to determine the impact of scientific papers and the attention received by the scientific community and the general public for each paper we traced altmetrics measures. Altmetrics, meant as a subset of scientometrics, firstly first introduced by Priem et al. (Priem 2012), is the study and use of scholarly impact measures based on activity in online tools and environments. The term has also been used to describe the metrics themselves and includes also non-traditional research impact measures. Altmetrics measurement derives from the social web and is increasingly used as an early indicator of research impact (cf. Section 1). The sources used for altmetrics are heterogeneous and include - beside traditional citations in peer-reviewed papers - mentions and citations in blogs, Wikipedia, Twitter or Facebook or reader counts on social reference managers and bookmarking platforms.

In this study we use the altmetrics provided by Plum Analytics^2^ (PlumX) which is one of the leading platforms that provides altmetrics (Nuzzolese 2019). It is a provider of alternative metrics created in 2012 and covers more than 52.6M of artifacts, metrics and sources of metrics that are collected into five separate categories: (i) **Citations:** contain both traditional citation indexes such as Scopus, as well as citations that help indicate societal impact such as Clinical or Policy Citations. (ii) **Mentions**: measures activities such as news articles or blog posts about research. Mentions is a way to tell that people are truly engaging with the research (examples are blog posts, comments, reviews, Wikipedia links, and news media); (iii) **Social Media interactions**: includes tweets, Facebook likes, etc. that reference the research. Social Media can help measure attention. Social media can also be a good measure of how well a particular piece of research has been promoted; (iv) **Captures**: indicate that someone wants to come back to the work. Captures can be a leading indicator of future citations (examples are bookmarks, code forks, favorites, readers, and watchers); (v) **Usage**: a signal if anyone is reading an article or otherwise using a research.

All the papers selected in the first stage of this study (i.e. during scientific literature review) have been collected by using their corresponding DOIs as the key for querying PlumX, as reported in Boetto et al. 2020. For each paper the citation count, the number of mentions on social media, the number of visits and clicks on online platforms, the number of readers on academic social networks (e.g. Mendeley), and the mentions on blogs, wikis and traditional media/press were collected (Table 1).

**Table 1.** The categories provided by PlumX along with an explanation about their semantics.

| Category | Explanation |
| --- | --- |
| Citations | Traditional citation indexes such as Scopus, and citations that help indicate societal impact such as Clinical or Policy Citations. |
| Mentions | Number of mentions retrieved in news articles or blog posts about research. |
| Social Media interactions | The number of mentions included in tweets, Facebook likes, etc. that reference a research work. |
| Captures | An indication that someone wants to come back to the work. |
| Usage | A signal that anyone is reading an article or otherwise using a research. |

This allowed us to calculate, for each paper, the above reported five metric categories (citations, captures, mentions, social media, and usage). Subsequently, given that the different categories have a different weight, as explained in Boetto et al. 2020, a Comprehensive Impact Score (CIS) was calculated. In fact, each paper has different numbers for each category considered and a standardized measure is needed to fairly quantify the communicative impact. CIS represents a unifying score over heterogeneous bibliometric indicators and categories. After CIS was computed for all the retrieved papers we used the z-score for obtaining standard values and the arithmetic mean for the average. Intuitively, the z-score is a numerical measure that gives us an idea of how far from the mean a data point is. Hence, a z-score is a scalar value that can be positive (i.e. the score is above the mean) or negative (i.e. is below the mean). Finally, we computed the quantiles of the resulting CIS values and identified a threshold t.

### 2.3 Data summary and synthesis

We report descriptive statistics related to the different metric categories of the selected papers (mean, standard deviation - SD, median, and confidence interval - CI). We considered the papers in the upper quantile (95%) for each metric category (citations, capture, mentions, social media and usage). For each category we also reported the value (e.g. number of citations or mentions) associated with the 95% quantile. Then, for papers above the identified CIS threshold, we described the main study characteristics in terms of publication date, publication type, aim of the study, and authors’ nationality.

## 3. Results

The search conducted yielded 442 potentially relevant papers. After deduplication and pertinence screening, 239 papers met the eligibility criteria for review and scientometric analysis (Figure 1).

**Figure 1.**
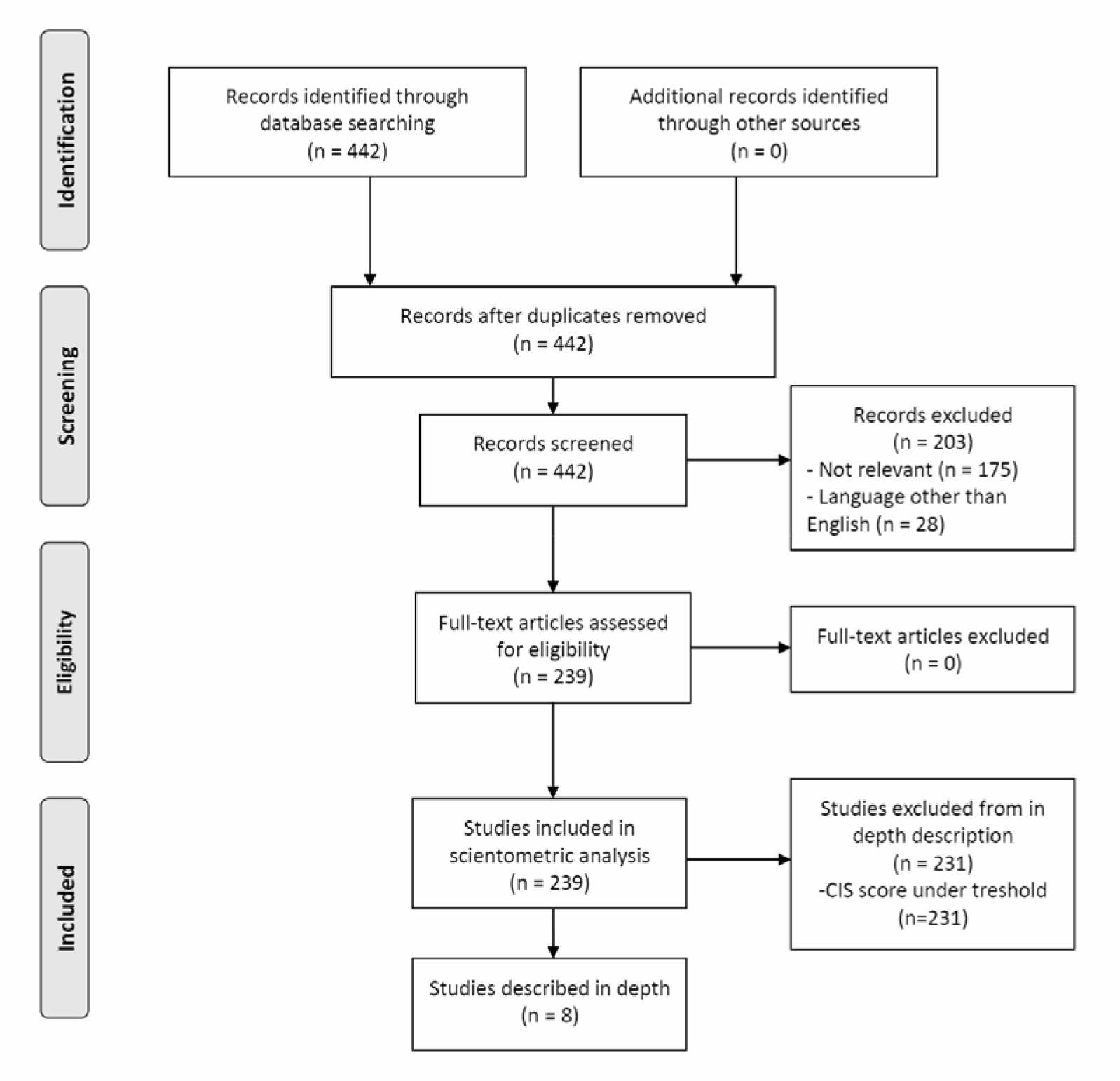
Scientific literature review flowchart (Prisma guidelines flowchart)

63.6% of the papers (152 out of 239) were editorials, commentaries or letters (mainly reported data). 10.5% of the papers (25 out of 239) were secondary papers, mainly narrative reviews, which collected the knowledge available up to that point on some specific topics (i.e. genomics of the virus, transmissibility, etc.). The remaining 25.9% (62 out of 239) were original primary studies: among these, case reports accounted for 42%, while in vitro or in vivo studies or genomic studies accounted for 21% of the total. The remaining primary studies were cohort studies, case control studies and surveys.

As reported in Table 2, the mean of “citations” for each paper resulted in 1.63 (median: 0; max: 82). The mean of “mentions” per paper in blogs and news was 10 while the mean of “social media” interactions resulted in 1,250 per paper. On average each paper had 1.69 “captures” and 0.07 “usages”, as defined in the Methods section.

**Table 2.** The statistics recorded for metric categories including the citation count.

| CATEGORY | METRIC | MAX | MEAN | MEDIAN | CATEGORY | METRIC | MAX | MEAN | MEDIAN |
| --- | --- | --- | --- | --- | --- | --- | --- | --- | --- |
| CITATIONS | Citation Count | 82 | 1.63 | 0 | CAPTURES | Readers | 161 | 1.69 | 0 |
|  | TOTAL CITATIONS | 82 | 1.63 | 0 |  | TOTAL READERS | 161 | 1.69 | 0 |
| MENTIONS | Blog Mentions | 22 | 0.95 | 0 | USAGE | Abstract Views | 15 | 0.07 | 0 |
|  | News Mentions | 253 | 8.95 | 0 |  | TOTAL USAGE | 15 | 0.07 | 0 |
|  | Q&A Site Mentions | 3 | 0.05 | 0 |  |  |  |  |  |
|  | References | 4 | 0.06 | 0 |  |  |  |  |  |
|  | TOTAL MENTIONS | 277 | 10.00 | 0 |  |  |  |  |  |
| SOCIAL MEDIA INTERACTIONS | Shares, Likes & Comments | 33043 | 582.06 | 0 |  |  |  |  |  |
|  | Tweets | 14409 | 457.36 | 29.5 |  |  |  |  |  |
|  | TOTAL SOCIAL MEDIA | 45,197 | 1,250.34 | 36.5 |  |  |  |  |  |

The papers positioned in the upper quantile (95%) for each category considered are reported in Table 1 in the Supplementary materials.

For citations, the papers in the 95% quantile are [Supplementary materials, Table 1 IDs: 1, 2, 3, 4, 5, 6, 7, 8, 9, 10, 11]. The 3 papers with the highest number of citations resulted “*Clinical features of patients infected with 2019 novel coronavirus in Wuhan, China*” by Huang C. et al. with 82 citations (Huang 2020), “*A familial cluster of pneumonia associated with the 2019 novel coronavirus indicating person-to-person transmission: a study of a family cluster*” by Chan J. et al. with 71 citations (Chan 2020) and “*Epidemiological and clinical characteristics of 99 cases of 2019 novel coronavirus pneumonia in Wuhan, China: a descriptive study*” by Chen N. et al. with 27 citations (Chen 2020).

For mentions, the papers in the 95% quantile are [Supplementary materials, Table 1 IDs: 1, 2, 3, 8, 9, 12, 13, 14, 15, 16, 17]. The 3 papers with the highest numbers of mentions resulted: “*Clinical features of patients infected with 2019 novel coronavirus in Wuhan, China*” by Huang C. et al. with 277 mentions (Huang 2020), which is also the paper with the highest number of citations, “*Homologous recombination within the spike glycoprotein of the newly identified coronavirus may boost cross*lJ*species transmission from snake to human*” by Wei J. et al. with 203 mentions (Wei 2020), and “*Transmission of 2019-nCoV Infection from an Asymptomatic Contact in Germany*” by Rothe C. et al with 154 mentions (Rothe 2020).

For social media interactions, the papers in the 95% quantile are [Supplementary materials, Table 1 IDs: 1, 2, 3, 7, 8, 9, 12, 14, 15, 16, 18]. The paper with the highest numbers of social media interactions resulted again “*Clinical features of patients infected with 2019 novel coronavirus in Wuhan, China*” by Huang C. et al. with 45197 social media interactions (Huang 2020), followed by “*First Case of 2019 Novel Coronavirus in the United States*” by Michelle L. Holshue et al. with 21.627 social media interactions (Holshue 2020), and “*Epidemiological and clinical characteristics of 99 cases of 2019 novel coronavirus pneumonia in Wuhan, China: a descriptive study*” by Chen N. et al. with 20.758 social media mentions (Chen 2020).

Considering captures a leading indicator of future citations, the papers in the upper quantile are [Supplementary materials, Table 1 IDs: 5, 6, 10, 12, 19, 20, 21, 22, 23, 24, 25]. The top 3 papers resulted “*The continuing 2019-nCoV epidemic threat of novel coronaviruses to global health - The latest 2019 novel coronavirus outbreak in Wuhan, China*” by Hui DS et al. with 161 captures (Hui 2020), “*Outbreak of pneumonia of unknown etiology in Wuhan, China: The mystery and the miracle*” by Lu H. et al. with 58 captures (Lu 2020), and “*Full-genome evolutionary analysis of the novel coronavirus* (*2019-nCoV*) *rejects the hypothesis of emergence as a result of a recent recombination event*” by Paraskevis D. et al. with 22 captures (Pareskevis 2020).

For usage, which represents the number of abstract views, only one paper was relevant [ID: 16]: “*First Case of 2019 Novel Coronavirus in the United States*” by Michelle L. Holshue et al. with 15 usages (Holshue 2020).

To obtain a unique and omni-comprehensive metric score we calculated the CIS. The association of papers to their corresponding CIS values is published in a spreadsheet, which is available online^3^. The following statistics provide a summary of the recorded CIS values (Figure 2): max=5.21, min=-0.21, mean=0.02, median=-0.19. The threshold identified is t=1.04. Such a threshold is the value of the 95% quantile and allows us to record 8 out of 239 papers (Figure 1, Figure 2) as potentially more impactful. Lower values for t are not significant for capturing relevant works (as explained in Boetto et al. 2020). The resulting 8 most impactful studies’ main features are described in Table 3. Among those, 6 papers are case reports, 1 methodological study, 1 editorial. First authors come from China (n = 6), USA (n=1) and Germany (n=1). The 8 papers’ main topics are: case/s description (n=5), outbreak investigation (n=2) and 1 genomic study.

**Figure 2.**
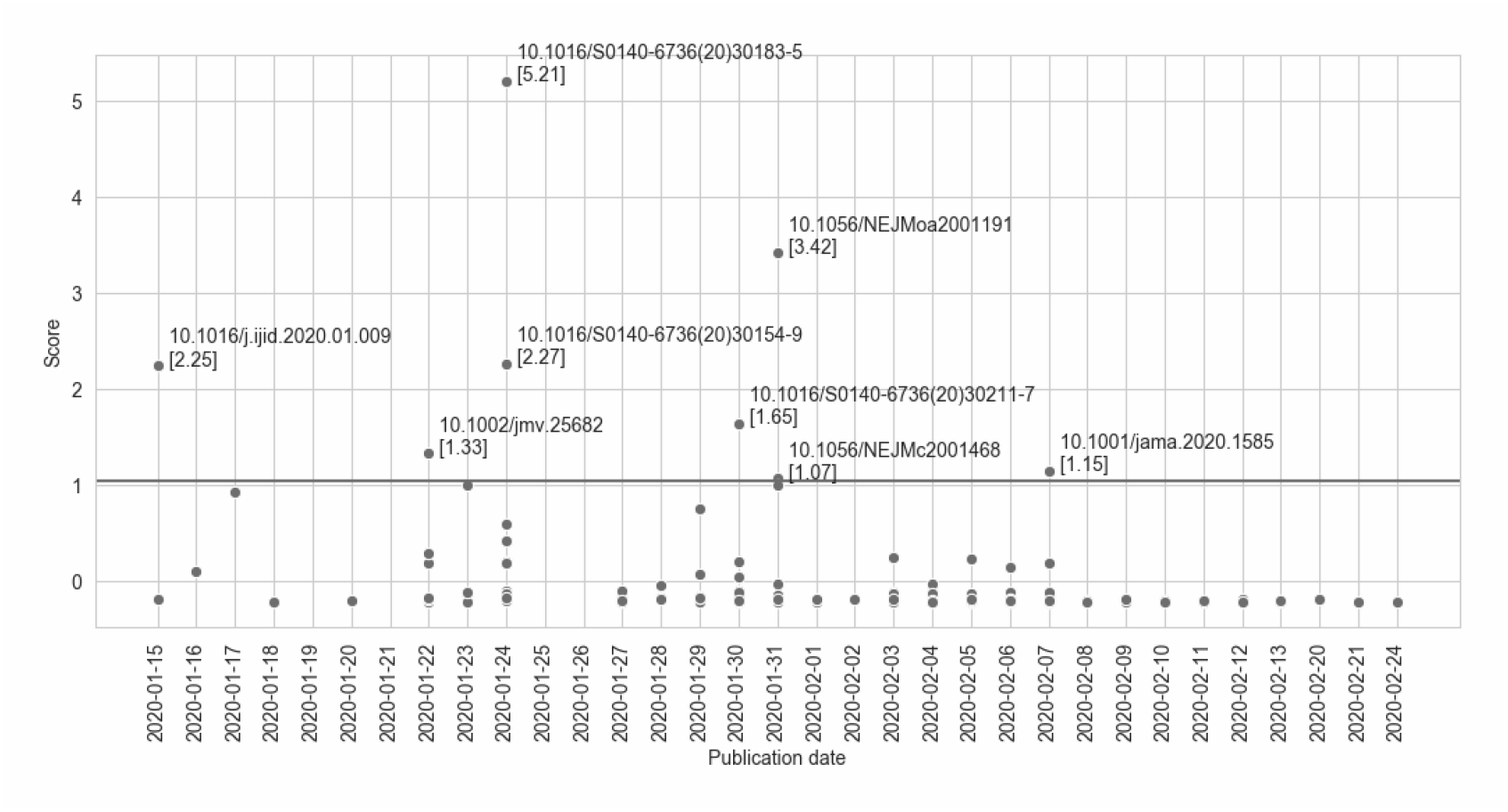
Scientific papers’ CIS score. The horizontal line represents the defined score threshold.

In particular, the 3 articles with higher CIS were “*Clinical features of patients infected with 2019 novel coronavirus in Wuhan, China*” (Huang 2020), “*First Case of 2019 Novel Coronavirus in the United States*” (Holshue 2020), and “*A familial cluster of pneumonia associated with the 2019 novel coronavirus indicating person-to-person transmission: a study of a family cluster*” (Chan 2020), as reported in Table 3. The three papers - 1 cohort study and 2 case studies - were all published in high impact journals (*The Lancet* and the *New England Journal of Medicine*).

The first paper, by Chaolin Huang et al. (Huang 2020), was published online on February 15th 2020 and reports a cohort of 41 patients with laboratory-confirmed SARS-CoV-2 infection. Patients had serious, sometimes fatal, pneumonia and were admitted to the designated hospital in Wuhan, China, by Jan 2, 2020. The study shows that the time between hospital admission and ARDS (Acute Respiratory Distress Syndrome) was as short as 2 days and that at this stage the mortality rate was high (15%). Also, the Authors recommend that faecal and urine samples should be tested to exclude a potential alternative route of transmission.

The second paper, by Michelle L. Holshue et al. (Holshue 2020), was published online on January 31th 2020 and reports the clinical features of the first reported patient with SARS-CoV-2 infection in the United States. The Authors describe key aspects of the case including the decision made by the patient to seek medical attention after reading public health warnings about the outbreak, the identification of possible SARS-CoV-2 infection, which allowed for prompt isolation of the patient and subsequent laboratory confirmation of COVID-19, as well as for admission of the patient for further evaluation and management.

The third paper with the highest comprehensive impact score is the study by Chan J. et al. (Chan 2020) in which the Authors report the epidemiological, clinical, laboratory, radiological, and microbiological findings of five patients in a family cluster who presented with unexplained pneumonia after returning to Shenzhen, Guangdong province, China, after a visit to Wuhan, and an additional family member who did not travel to Wuhan.

## 4. Discussion

The aim of this study was to describe and quantify the impact of early scientific production in response to the COVID-19 pandemic. In an increasingly connected world, tracing the traditional and non-traditional metrics measures of scientific papers can help to understand and evaluate their communicative impact on the researchers’ community and general population.

The COVID-19 pandemic is taking place in an historical period characterized by high rapidity of communications, through traditional media, internet, and social networks. At the present, the levels of digitalization and the speed of data and information exchange at a global level are incredibly higher than in any previous epidemics and pandemics (e.g. SARS in 2003, H1N1 in 2009).

It is therefore of primary importance to quickly identify the most relevant information, data and scientific evidence because this can be useful to guide policymakers, healthcare professionals and the general population in a time of crisis. When there is no consistent scientific data nor strong evidence, it is particularly relevant to identify which scientific information is capturing the attention. This can be done using several alternative bibliometric tools (i.e. altmetrics) that can be useful for tracing which scientific papers bounce more and have a greater impact, particularly in the early phases of an epidemic, when traditional metrics (e.g. citations) may not be as timely, relevant or exhaustive.

From our analysis, the three papers with the highest number of citations are those with the greatest impact on the scientific community. In fact, all of them are original studies with data from “the field”, describing the clinical characteristics, the clinical course and the transmission routes of COVID-19 cases.

“Mentions” are the number of mentions retrieved in news or blog posts. Except for one paper on the clinical characteristics of COVID-19 patients in the first Chinese outbreak (the same with the highest number of citations), it is interesting to note that the other two most mentioned papers describe the first cases of SARS-CoV-2 infection in Europe (Germany) and the United States. Clearly those are interesting aspects for a wider audience than the scientific community alone.

The “social media” interactions are consistent with the dynamics of traditional citations and mentions. The papers about the first cases outside China (especially in the United States) resonate the most, alongside the studies that describe the COVID-19 clinical manifestations.

“Captures” is an alternative metric indicating that someone wants to go back to the paper or wants to deepen the topic. From our results and in our opinion, the most “captured” papers are those with appealing titles using evocative words, for example “*The continuing 2019-nCoV* ***epidemic threat*** *of novel coronaviruses to* ***global health*** *- The latest 2019 novel coronavirus outbreak in Wuhan, China*” (Hui 2020) or “*Outbreak of pneumonia of* ***unknown etiology*** *in Wuhan, China:* ***The mystery and the miracle***” (Lu 2020). The first two papers are commentaries, while the third one is a genomic study on the origin of SARS-CoV-2.

With the CIS we unify and identify what paper is showing more attention both from the scientific community and general public. This is because the CIS in a standardized way, beside citations also catches the impact of other traditional media and social networks.

Again, the paper with highest CIS resulted “*Clinical features of patients infected with 2019 novel coronavirus in Wuhan, China*” by Chaolin Huang et al. (Huang 2020) and published on *The Lancet*, which rated first also in citations and mentions. Intuitively, this is the paper with the greatest impact because it is the first (January 24th 2020) describing a population of patients affected by the new disease, identifying its epidemiological, clinical, laboratory, and radiological characteristics and treatment and clinical outcomes. These data and information are clearly of great interest to both the scientific community and the general population.

Scientific citations, newspapers and blogs usually report more reliable information, whereas the most “sensational” things, with less technical details and a more appealing presentation - even within scientific works - bounce back on social media, as expected. As recently reported in this viewpoint (Merchant 2020), it is therefore mandatory for scientific communication to be effective not only for professionals but also for the general public. This is crucial for counteracting misinformation which is regarded as a real threat for public health and to promote information exchange that could facilitate any public health intervention. In fact, sharing the best possible scientific evidence is crucial for systems’ preparedness, as well as it is also essential to monitor social media to avoid misinformation.

In conclusion, being able to monitor how communications spread from the scientific world toward the general population, using both traditional and alternative metric measures, seems extremely important, especially in the early stages of a pandemic.

## Data Availability

Available upon request to the Authors

## Supplementary data

Supplementary data are available at Research Evaluation Journal online.

## Footnotes

1 The New York Times (2020). ‘W.H.O. Fights a Pandemic Besides Coronavirus: An ‘Infodemic’.’. https://www.nytimes.com/2020/02/06/health/coronavirus-misinformation-social-media.html

2 https://plumanalytics.com/

3 http://bit.ly/2T7Mdu4

